# Item generation for a new patient-reported outcome measure: The non-traumatic anterior knee pain (AKP)-YOUTH scale

**DOI:** 10.1101/2023.12.06.23299599

**Authors:** Marie Germund Nielsen, Kristian Damgaard Lyng, Sinead Holden, Simon Kristoffer Johansen, Marinus Winters, Michael Skovdal Rathleff

**Author notes:** **Corresponding Author** Kristian Damgaard Lyng, Department of Health Science and Technology, Faculty of Medicine, Aalborg University, Selma Lagerlöfs Vej 249, 9260 Gistrup, Denmark. Shared first authorship (Marie Germund Nielsen, Kristian Damgaard Lyng). All authors have read and approved this manuscript. **Competing interests** None stated. **Data availability statement** Researchers interested in the data from the AKP-Youth scale study may contact the principal investigator, Prof. Michael Skovdal Rahtleff,.

## Abstract

**Question:** Which domains are important to develop a preliminary item bank for a new patient-reported outcome measure relating to adolescents with non-traumatic anterior knee pain?

**Design:** Multiple methods: semi-structured interviews,

**Participants:** Twenty-one adolescents with anterior knee pain participated in semi-structured interviews which explored their experience of living with knee pain. Following thematic analysis, we generated an item bank based on the domains which emerged from the impact their knee pain had on their daily life. Ten clinical experts provided input on the preliminary item bank via an online survey. Cognitive interviews were conducted using the think-aloud approach with ten adolescents to evaluate the comprehensibility and face validity of the items.

**Results:** From the interviews we identified four overarching domains where adolescents were impacted by their knee pain: knee symptoms, limitations in physical activity/sport, limitations in social activities, and emotional impact of pain. Eighteen items was initially developed and expanded to 23 following clinical expert input. The cognitive interviews with adolescents demonstrated that the items were comprehensive, understandable, and relevant for adolescents.

**Conclusion:** This study developed an item bank of 23 items. These spanned four domains of impact for adolescents with anterior knee pain. The items had good face validity and were deemed relevant and understandable for adolescents with knee pain. Further steps are needed to validate and reduce the items for the non-traumatic anterior knee pain (AKP)-YOUTH scale.

## Introduction

Musculoskeletal pain is common in adolescents, affecting almost one in three (1,2). Knee pain is one of the most common pain types (3,4) and is associated with disability, low quality of life, and difficulties participating in physical activity, sport, and social activities (5–7). Several painful conditions are included under the definition of non-traumatic anterior knee pain. The most common conditions are Osgood Schlatter disease, patellofemoral pain, and Sinding-Larsen Johansson, which are associated with pain and functional impairments which often span months or even years (7–11).

Patient-reported outcomes are reports of patients’ health conditions coming directly from the patients and are important to gain patients’ perspectives on their symptoms and condition (12). Patient-reported outcome measures (PROMs) can either be generic (e.g., measures of quality of life that are designed for use across multiple conditions) or condition-specific PROMs, which are designed to capture changes in a specific condition (e.g., Knee Injury and Osteoarthritis Outcome Score (KOOS) (13)). No PROMs have been developed specifically for adolescents with non-traumatic anterior knee pain.

The PROMs currently used in clinical research on non-traumatic knee pain in adolescents (e.g., KOOS, KOOS-Child and Pedi-IKDC) are not specifically developed for adolescents with non-traumatic anterior knee pain (13–15). These PROMs may not adequately capture the adolescent’s experience and creates uncertainty in what is measured (16,17). This can lead to inaccuracies when estimating the effect of an intervention or the prognosis of the knee condition (18). The main objectives of this study were to identify domains and develop a preliminary item bank for adolescents with non-traumatic knee pain. This item bank should later contribute to the non-traumatic anterior knee pain (AKP)-YOUTH scale.

## Materials and methods

### Design

To identify domains of relevance for the AKP-YOUTH scale, we incorporated lived experiences from adolescents with anterior knee pain, clinical experts, and research on anterior knee pain which was mapped to the International Classification of Functioning, Disability and Health (ICF) model. The development consisted of the following steps (Figure 1):

**Figure 1:**
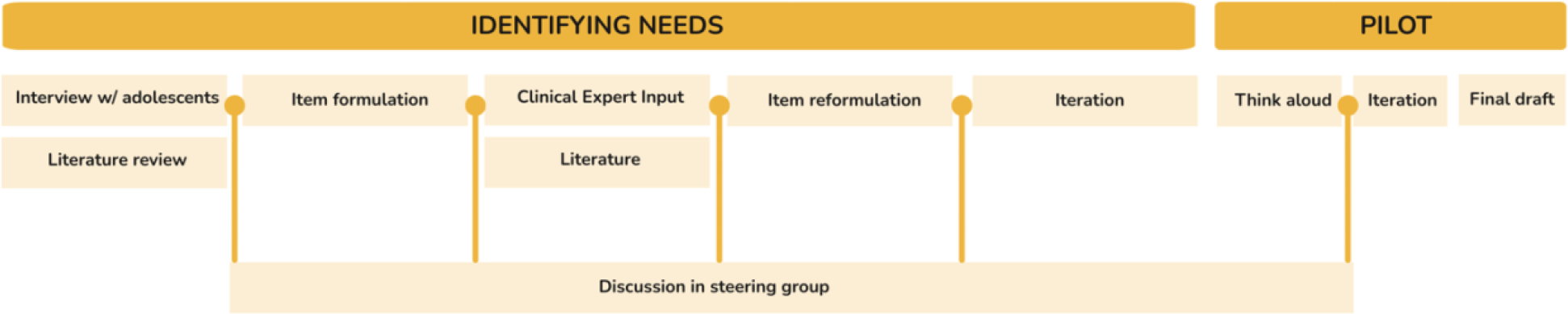
DEVELOPMENT PROCESS OF THE AKP-YOUTH PROM.

1. Identification of domains of impact for adolescents with knee pain through semi-structured interviews with adolescents
2. Preliminary item bank formulation
3. Clinical expert input on identified domains and preliminary item bank
4. Cognitive interviews of adolescents using the think-aloud approach.
5. Translation of content

Data were processed and stored in accordance with the regulations of the General Data Protection Regulation (GDPR). Ethical approval by the local committee was applied for but was deemed exempt from full approval due to the low-risk nature of the study. Written informed consent was obtained from the participants (15 years of age or older) or their legal guardian(s) (if below 15 years).

### Step 1: Identification of relevant domains through interviews

To ensure domains included in an item bank reflected the lived experience of adolescents with non-traumatic anterior knee pain (content validity), we conducted qualitative semi-structured interviews with adolescents. An interview guide was designed by members of the steering group (Appendix 1) based on existing research, members experience with this patient population, and the ICF model (19). It consisted of open-ended questions, covering different aspects of adolescents’ experience of living with knee pain, while exploring which parts of the experience the adolescents deemed important to improve. The interview guide was piloted on three adolescents with knee pain to ensure comprehensibility and incited deep descriptions from participants (20). The interview guide was iterated as new themes emerged from the interviews. Adolescents aged 10 to 19 years with any type of non-traumatic anterior knee pain for over three months were eligible for inclusion in the interviews. Adolescents with a traumatic origin of pain or any other origin of pain (e.g., systemic disease, tumour) and those who were not fluent in Danish were ineligible. McConaughey’s recommendations for interviewing adolescents were used to ensure all necessary precautions were taken (21). The interviews lasted between 30 and 60 minutes. The interviews were conducted in person (n = 15) or over Microsoft Teams (n = 6). Parents were invited to be present during the interviews. All interviews were audio recorded and transcribed verbatim by the interviewer and two student assistants using NVivo 12 (NVivo qualitative data analysis software; QSR International Pty Ltd. Version 12, 2018). Data were analysed using the thematic text analysis approach described by Braun and Clarke (24), which included the following steps: familiarisation, coding, searching for themes, reviewing themes, defining, and naming themes, and writing up themes into a narrative form. To familiarise themselves with the raw data, the transcripts of the interviews were read multiple times by two coders. From this thematic coding, potential themes were categorized by one researcher via inductive interpretation. All potential themes were summarised, combined, and presented to the rest of the steering group as potential domains and items. All themes were discussed, reviewed, and refined by the project group. As new themes were identified, the transcripts were reread to ensure the themes and their related domains were empirically grounded.

### Step 2: Preliminary item bank formulation based on identified domains of impact

Based on the results and identified themes from the interviews in step 1 and knowledge from the literature (8,22–24) on adolescent anterior knee pain, initial items were formulated using guidelines from Streiner et al (25). From the literature we used phrasing of items and response options from other questionnaires in the preliminary item bank (14,15,26). This was conducted by the study’s steering group. The following criteria were used to generate preliminary items; the items should:

1. Be discriminatory; measure differences within the construct, important to the adolescent with non-traumatic anterior knee pain
2. Be linked to adolescents’ knee condition
3. Be able to measure changes over time
4. Measure a present state (i.e., does not say something about what will happen/is expected to happen in the future)
5. Be applicable/relevant to all (>95%); all possible adolescents with knee pain should be able to score on the item (e.g., an item on playing football may not be appropriate as not all adolescents with knee pain engage in football regularly).
6. Each domain should include one construct

Preliminary items were as simple as possible (i.e., simplicity in phrasing, no jargon, no ambiguity, no double-barrelled questions, and no value-laden words) to ensure understandability regardless of health literacy (25). The formulation of the preliminary items was an iterative process between the members of the steering group and was repeated until agreement. All decisions and changes to the items were discussed collaboratively within the steering group, which consisted of the authors who had clinical and research experience with anterior knee pain among adolescents.

### Step 3: Clinical expert input

We aimed to incorporate input from clinical experts into the identification of the domains and preliminary items to ensure the item bank covered the important domains for this population. Surveys were conducted in English, developed, and sent through Research Electronic Data Capture tools (REDCap™) hosted at Aalborg University (27,28). Some of the items, were created in Danish, and hence the steering group translated the items into English for the purpose of the survey. The participants were asked if the domains and items were adequately comprehensive. Participants were also asked if the item bank missed any domains, or items within domains. Participants were eligible to participate as experts if they had a minimum of 5 years of experience with adolescents with knee pain and had been involved in research on knee pain (29). No domains or items were removed as this step was to provide input on phrasing and to identify any potential domains or items of relevance that had not yet been captured by the initial set of items.

### Step 4: Think-aloud exercise with adolescents

A think-aloud exercise was conducted with a researcher acting as ‘narrator’ (KDL) and included the assessment of setup, usability, comprehensibility, and face validity via reflection-in-action (30). To ensure that the wording of the item bank were applicable and readable to as many 10-19 years as possible, we expanded the recruitment to include children and adolescents between 8-19 years. The think-aloud exercise was conducted online via Microsoft Teams using a process guide. Audio and video from the interviews were recorded (see Appendix 2 for think-aloud interview process) (31). Parents were allowed to sit next to the adolescent to replicate a situation in which adolescents would complete it with parents present. The adolescents received a link to the item bank and were asked to print it out prior to the think-aloud exercise. During the think-aloud exercise, the participants were asked to complete the items while reading and thinking out loud about the items and responses. The participants were instructed to say whatever came to their minds as they went through the items. This allowed the identification of any difficulties in understanding phrases or concepts. If the adolescents would stop or pause, the researcher would prompt (e.g., ‘why did you pause on this’) them to articulate their reflections while reading the items to identify any misunderstanding or confusion of each item. During all think-aloud exercises, the researcher collected field notes which were updated by reviewing the video recordings after the exercise to ensure nothing was overlooked (30). Sampling of participants was continued until saturation. We adjusted the items according to the responses on an ongoing basis. And transcripts from the think-aloud exercise were imported into Microsoft Word, analysed, and presented to the rest of the steering group for final interpretation. Reiterations of the item bank were discussed within the steering group and decisions were made by consensus.

### Step 5: Translation of content

All Danish content was translated through a dual-panel approach (32). The first panel consisted of the author group, consisting of five individuals (two physiotherapist, one nurse, one exercise scientist, one information specialist) fluent in Danish and English. Five were native Danish speakers, and one was a native English speaker. All independently translated the Danish all study-derived content into English. The aim of their translation where to create translations that could be understood by the target population (adolescents with non-traumatic knee pain aged between 10-19 years old). The panel then synthesized and compared their translations with linguistic imprecisions, language- and cultural appropriation in mind. Through consensus, a final version was derived. Hereafter, a new panel consisting of four laypersons were established and this panel were instructed to review the final translation from the first panel. The second panel provided inputs to re-wording or re-phrasing of the translation. In cases where the item bank and responses were derived from other validated PROMS, no translation were performed, and the original formulation was kept. All translated material is presented in Appendix 8.

## RESULTS

### Step 1: Identification of domains of relevance for adolescents with knee pain through semi-structured interviews with adolescents

Twenty-one adolescents with non-traumatic knee pain participated in the interviews to give their experience of living with knee pain. Detailed demographics are presented in Table 1. The thematic analysis revealed 12 domains. Summaries of each domain together with illustrative quotes, were discussed within the steering group and distilled into four overarching domains of importance. These were symptoms, limitations of physical activity, limitations in social activity, and the emotional impact of pain.

**Table 1:**
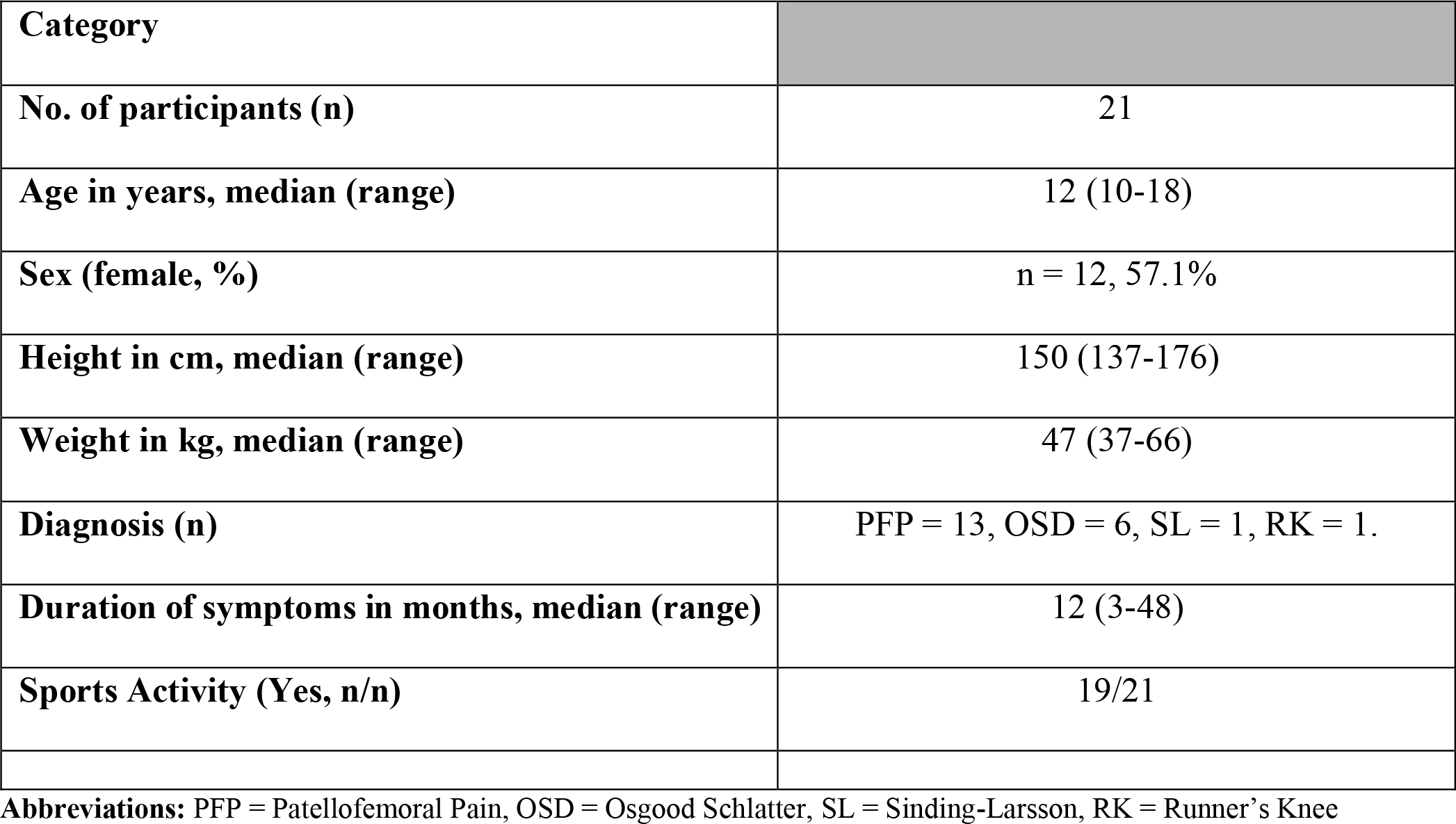
DEMOGRAPHICS OF PARTICIPANTS IN STEP 1.

| Category |  |
| --- | --- |
| No. of participants (n) | 21 |
| Age in years, median (range) | 12 (10-18) |
| Sex (female, %) | n = 12, 57.1% |
| Height in cm, median (range) | 150 (137-176) |
| Weight in kg, median (range) | 47 (37-66) |
| Diagnosis (n) | PFP = 13, OSD = 6, SL = 1, RK = 1. |
| Duration of symptoms in months, median (range) | 12 (3-48) |
| Sports Activity (Yes, n/n) | 19/21 |

**Abbreviations:** PFP = Patellofemoral Pain, OSD = Osgood Schlatter, SL = Sinding-Larsson, RK = Runner's Knee

### Step 2: Preliminary item bank formulation based on expert (steering group)

The steering group identified four key domains (symptoms, limitations in physical activity, limitations social activity and emotional impact of pain). A description of the topics after this stage is included in Appendix 3. Also, existing literature was used and where possible, phrasing or structure of phrasing of items and response options from exciting questionnaires. Preliminary item formulations are included in Appendix 4.

### Step 3: Clinical expert input

Eleven clinical experts responded to the online questionnaire (demographics and clinical experience are outlined in Table 2). The expert origins from the following countries: Australia 1(9.1%), Denmark 3(27.3%), Ireland 1(9.1%), United Kingdom 3(27.3%) and USA 1(9.1%). Overall, the experts deemed the identified domains and preliminary items relevant and comprehensive for adolescents with anterior knee pain. The experts made suggestions for improvements in the phrasing of the included items. Some of the feedback pertained to overlap in items. Based on the clinical expert input, six items were added (Appendix 5). This was done to better capture the domain of physical activity and exercise (3 items) to better capture the general ability to be active and how this is influenced by knee pain. This was triangulated with the themes identified in the interviews with adolescents and based on this, additional items were added to ensure full comprehensiveness in the item bank. In addition, items were added for impact on sleep (symptoms domain), standing from prolonged sitting (limitations in activity), and difficulty kneeling (limitations in activity). The items added are listed in appendix 5, and they correspond to items 3, 4, 5, 6, 8, and 13 in the questionnaire.

**Table 2:**
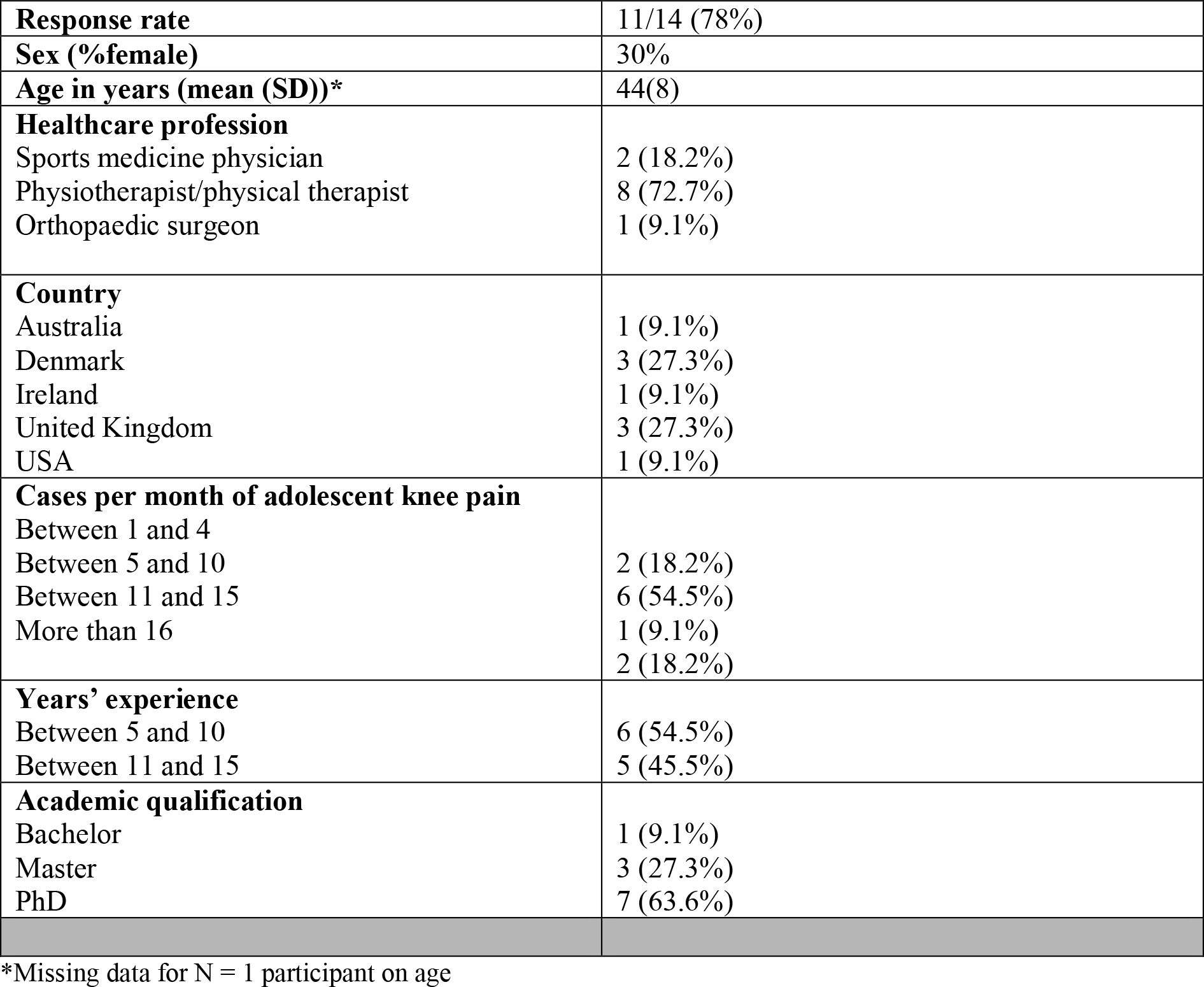
DEMOGRAPHICS OF PARTICIPANTS FOR STEP 3.

### Step 4: Think-aloud exercise with adolescents

Cognitive interviews were undertaken on the final 23 items in a draft questionnaire (33). Ten adolescents (8 with non-traumatic anterior knee pain, and 2 were pain-free) participated in the think-aloud exercise. Demographics are presented in Table 3. Themes generated from think-aloud are presented in table 4. All participants were able to read and understand the formulations and response categories used in the think-aloud exercise. In the analysis we identified issues and incorporated these. In the first theme, *usabilit*y, the adolescents expressed that REDCap was easy to use and only required little to no extra mental effort to navigate through the items. One participant highlighted the potential need for a read-aloud function for those with a reading-disability. Within the theme of *visibility*, the adolescents expressed that it was easy to recognise the intended meaning of the items and that the instructions on how to reply were clear. In the *understandability* theme, the adolescents highlighted several ideas for rephrasing items questions (see Appendix 6 for item formulation after think-aloud). Furthermore, most of the participants expressed difficulty in understanding the difference between severity and frequency related to two items. Despite a few proposed corrections from the participants, the adolescents expressed that the overall reading flow was good but highlighted that the flow could be improved if some of the questions were shortened. No action was taken as the overall impression was good and in general, all participants expressed that the content was both accurate and appropriate in terms of their own lived experience, and thereby the item list was considered acceptable and comprehensive.

**Table 3:** DEMOGRAPHICS OF PARTICIPANTS STEP 4.

| Category |  |
| --- | --- |
| No. of participants (n) | 10 |
| Age in years, median (range) | 11.5 (8-17) |
| Sex (female, %) | n = 6, 60% |
| Height in cm, median (range) | 151.5 (140-178) |
| Weight in kg, median (range) | 41.5 (27-62) |
| Diagnosis (n) | 6 = OSD, 2 = PFP, 2 = pain-free |
| Sports Activity (Yes, n/n) | 10/10 |

**Abbreviations:** PFP = Patellofemoral Pain, OSD = Osgood Schlatter,

**TABLE 4:** THEMES GENERATED FROM THINK-ALOUD.

| **Theme** | **Description** |
| --- | --- |
| *Usability* | Practical use of the items, including time, navigation, clicks and mental effort. |
| *Visibility* | Level of ease to recognize the key messages and instructions given in the questionnaire |
| *Content* | Accuracy and appropriateness of content included in the questionnaire |
| *Understandability* | Understandability and meaning of words used in the questionnaire |
| *Flow* | Ability to navigate through each category and general reading flow of questionnaire |
| *Acceptability* | General accept of the relevancy of the overall questionnaire |

## DISCUSSION

We identified a preliminary set of items to be used in a condition-specific PROM for adolescents with non-traumatic knee pain. We combined multiple steps to ensure that the domains identified were of importance to adolescents, clinicians, and researchers. Our results from the cognitive interviews showed that the preliminary set of items had good face and content validity. The clinicians’ additions to the domains emerged from a clinical rationale which are valuable for clinicians using the tool for treating adolescents. Several clinical trials have used PROMs for non-traumatic anterior knee pain (22,34) however none of them have been developed specifically for this population. The most frequently used PROMs are the KOOS and KOOS Child, the Anterior Knee Pain Scale, and the Pedi-IKDC (13–15,26). The KOOS child covers five domains: pain, other symptoms, function in daily living (ADL), function in sport and recreation (Sport/Rec), and knee-related quality of life (QOL) (14). The Anterior Knee Pain Scale covers pain, symptoms, and knee function domains, while the Pedi-IKDC covers three domains: symptoms, sports activity, and knee function (15,26). None of these PROMs were originally developed based on interviews with adolescents suffering from non-traumatic anterior knee pain. These PROMs are either physician-based, includes symptoms items not relevant to our population or developed without patient involvement. Our interviews with 21 adolescents revealed two domains of importance that are not included in the currently used PROMs. The domains missing in current PROMs are limitations in social activities (e.g., ability to do things with your friends) and the emotional impact of the pain (e.g., how often you feel sad because of knee pain). This highlights the importance of engaging adolescents to ensure that PROMs captures domains of importance to them. Where possible, we used phrasing or structure of phrasing of items and response options from other questionnaires in the preliminary bank of items (14,15,26). This included frequency measures such as “Never”, “Rarely”, “Sometimes”, “Often”, and “Always”. Likert scales were used to assess the degree of difficulty such as “Not at all difficult”, “A little bit difficult”, “Somewhat difficult”, “Very difficult”, and “Extremely difficult”. The aim of the response options was to make answering easy and comprehensible for adolescents and make sure we used (where possible) phrasing tested multiple times with this age group.

### Meaning of findings

Our study introduces a new item bank for adolescence with non-traumatic anterior knee pain. Applying such items (and as a PROM in the future) can lead to better trials where researchers capture what is most important to the end-users. Previous cross-sectional research shows that Osgood Schlatter disease and Patellofemoral Pain (the two most common conditions of non-traumatic anterior knee pain have a significant impact on knee function, ADL, sports participation, quality of life, sleep, and emotional well-being in adolescents (8). However, these studies have been conducted using PROMs decided upon by researchers. Previous developed PROMs are not comprehensive and has not been proven relevant among adolescence. This means that we may not capture the full impact of these conditions on adolescents with non-traumatic knee pain. This has implications both for interpreting the results of trials as well as the design of trials to target domains of importance to the adolescents.

### Limitations

We conducted the initial interviews in step 1 with children and adolescents from Denmark. It is unclear if our findings would have been different if adolescents were from other countries where physical activity patterns may be different. As an example, problems with bicycling were identified as an important domain in our interviews. In countries where adolescents do not bike regularly, this question may be less relevant. It is crucial that future adaptations cover the cultural differences that might exist. It could have been a strength if we adolescents were also involved in the item development (e.g., through workshops). However, the content validity for adolescents is ensured by the findings from the interviews and think-aloud exercises. We aimed to include children and adolescents with various forms of non-traumatic anterior knee pain to ensure that a potential future PROM would be suitable for all types of non-traumatic anterior knee pain between the age of 10 and 19 years of age. Therefore, it is unclear if there are domains and items specifically relevant for specific knee conditions within the broader group of adolescents with non-traumatic anterior knee pain. We conducted the think-aloud exercise online due to COVID-19 restrictions. This method made it easy and accessible for adolescents across Denmark to participate, but it is unclear if it affected the adolescents during the think-aloud exercise. We used a pragmatic Danish-English translation process, which might have affected our contributions/alterations to the items during the development. Three of the items (jumping, biking, running) may not be relevant to all adolescents. Future studies need to test the relevance of these items outside a Danish context.

### Implications and future research

Our preliminary item bank needs further testing in larger groups to test both validity, reliability and responsiveness before usage in research and clinical practice (35). Developing a valid PROM will provide health professionals understand the impact across domains of importance to adolescents and inform the treatment and progress of adolescents with non-traumatic anterior knee pain.

## Conclusion

A preliminary item bank was developed with inputs from adolescents with non-traumatic anterior knee pain, and clinical experts. The item bank revealed items involving biopsychosocial aspects and items not previously covered by other PROMs for adolescent knee pain. Further studies is needed to ensure that the most reliable and valid items has been identified prior to implementation in clinical practice and research.

## Data Availability

Researchers interested in the data from the AKP-Youth scale study may contact the principal investigator, Prof. Michael Skovdal Rahtleff, at.

## Acknowledgements

The authors would like to thank the expert clinicians, who provided valuable input to the development of the PROM. Additionally, the authors would like to thank all the participants including adolescents, parents, and clinicians who participated in the study. Finally, thank you to Rasmus Sloth Hoffman for his contribution to designing the study and acquisition of data.

## Declaration of generative AI in scientific writing

All authors declare that no artificial intelligent technology was used in any part of this study.

## References

1. Keeratisiroj O, Siritaratiwat W. Prevalence of self-reported musculoskeletal pain symptoms among school-age adolescents: age and sex differences. Scand J Pain. 2018;18(2):273–80.

2. Kamper SJ, Henschke N, Hestbaek L, Dunn KM, Williams CM. Musculoskeletal pain in children and adolescents. Braz J Phys Ther. 2016;20(AHEAD):0–0.

3. Rathleff MS, Roos EM, Olesen JL, Rasmussen S. High prevalence of daily and multi-site pain – a cross-sectional population-based study among 3000 Danish adolescents. Bmc Pediatr. 2013;13(1):191.

4. King S, Chambers CT, Huguet A, MacNevin RC, McGrath PJ, Parker L, et al. The epidemiology of chronic pain in children and adolescents revisited&colon; A systematic review. Pain. 2011;152(12):2729–38.

5. Rathleff MS, Rathleff CR, Olesen JL, Rasmussen S, Roos EM. Is Knee Pain During Adolescence a Self-limiting Condition? Am J Sports Medicine. 2016;44(5):1165–71.

6. Rathleff MS, Roos EM, Olesen JL, Rasmussen S, Arendt-Nielsen L. Lower Mechanical Pressure Pain Thresholds in Female Adolescents With Patellofemoral Pain Syndrome. J Orthop Sport Phys. 2013;43(6):414–21.

7. Guldhammer C, Rathleff MS, Jensen HP, Holden S. Long-term Prognosis and Impact of Osgood-Schlatter Disease 4 Years After Diagnosis: A Retrospective Study. Orthop J Sports Medicine. 2019;7(10):2325967119878136.

8. Rathleff MS, Winiarski L, Krommes K, Graven-Nielsen T, Hölmich P, Olesen JL, et al. Pain, Sports Participation, and Physical Function in Adolescents With Patellofemoral Pain and Osgood-Schlatter Disease: A Matched Cross-sectional Study. J Orthop Sport Phys. 2020;50(3):149–57.

9. Rathleff MS, Holden S, Straszek CL, Olesen JL, Jensen MB, Roos EM. Five-year prognosis and impact of adolescent knee pain: a prospective population-based cohort study of 504 adolescents in Denmark. Bmj Open. 2019;9(5):e024113.

10. Holden S, Olesen JL, Winiarski LM, Krommes K, Thorborg K, Hölmich P, et al. Is the Prognosis of Osgood-Schlatter Poorer Than Anticipated? A Prospective Cohort Study With 24-Month Follow-up. Orthop J Sports Medicine. 2021;9(8):23259671211022240.

11. Holden S, Roos EM, Straszek CL, Olesen JL, Jensen MB, Graven-Nielsen T, et al. Prognosis and transition of multi-site pain during the course of 5 years: Results of knee pain and function from a prospective cohort study among 756 adolescents. Plos One. 2021;16(5):e0250415.

12. Weldring T, Smith SMS. Patient-Reported Outcomes (PROs) and Patient-Reported Outcome Measures (PROMs). Heal Serv Insights. 2013;6:HSI.S11093.

13. Roos EM, Roos HP, Lohmander LS, Ekdahl C, Beynnon BD. Knee Injury and Osteoarthritis Outcome Score (KOOS)—Development of a Self-Administered Outcome Measure. J Orthop Sport Phys. 1998;28(2):88–96.

14. Örtqvist M, Roos EM, Broström EW, Janarv PM, Iversen MD. Development of the Knee Injury and Osteoarthritis Outcome Score for Children (KOOS-Child). Acta Orthop. 2012;83(6):666–73.

15. Nasreddine AY, Connell PL, Kalish LA, Nelson S, Iversen MD, Anderson AF, et al. The Pediatric International Knee Documentation Committee (Pedi-IKDC) Subjective Knee Evaluation Form: Normative Data. Am J Sports Medicine. 2017;45(3):527–34.

16. Anker SD, Agewall S, Borggrefe M, Calvert M, Caro JJ, Cowie MR, et al. The importance of patient-reported outcomes: a call for their comprehensive integration in cardiovascular clinical trials. Eur Heart J. 2014;35(30):2001–9.

17. Makrinioti H, Bush A, Griffiths C. What are patient-reported outcomes and why they are important: improving studies of preschool wheeze. Archives Dis Child - Educ Pract Ed. 2020;105(3):185–8.

18. Kluzek S, Dean B, Wartolowska KA. Patient-reported outcome measures (PROMs) as proof of treatment efficacy. Bmj Evidence-based Medicine. 2022;27(3):153–5.

19. International classification of functioning, disability, and health : ICF [Internet]. Version 1.0. Geneva : World Health Organization, [2001] ©2001; Available from: https://search.library.wisc.edu/catalog/999977181002121

20. Kvale S, Brinkmann S. Interviews: Learning the craft of qualitative research interviewing. 3. edition. Thousand Oaks, Calif.: Sage Publications; 2014. xviii, 405 pages p.

21. McConaughy S. Clinical interviews for children and adolescents : assessment to intervention. New York: Guilford Press; 2013. xv, 272 s. p.

22. Rathleff MS, Graven-Nielsen T, Hölmich P, Winiarski L, Krommes K, Holden S, et al. Activity Modification and Load Management of Adolescents With Patellofemoral Pain: A Prospective Intervention Study Including 151 Adolescents. Am J Sports Medicine. 2019;47(7):1629–37.

23. Winters M, Holden S, Lura CB, Welton NJ, Caldwell DM, Vicenzino BT, et al. Comparative effectiveness of treatments for patellofemoral pain: a living systematic review with network meta-analysis. Brit J Sport Med. 2021;55(7):369–77.

24. Johansen SK, Holden S, Pourbordbari N, Jensen MB, Thomsen JL, Rathleff MS. PAINSTORIES – Exploring the temporal developments in the challenges, barriers, and self-management needs of adolescents with longstanding knee pain: A qualitative, retrospective interview study with young adults experiencing knee pain since adolescence. J Pain. 2021;

25. Streiner DL, Norman GR, Cairney J. Health measurement scales: A practical guide to their development and use. 5th edition. Press OU, editor. Oxford University Press; 2015.

26. Kujala UM, Jaakkola LH, Koskinen SK, Taimela S, Hurme M, Nelimarkka O. Scoring of patellofemoral disorders. Arthrosc J Arthrosc Relat Surg. 1993;9(2):159–63.

27. Harris PA, Taylor R, Thielke R, Payne J, Gonzalez N, Conde JG. Research electronic data capture (REDCap)—A metadata-driven methodology and workflow process for providing translational research informatics support. J Biomed Inform. 2009;42(2):377–81.

28. Harris PA, Taylor R, Minor BL, Elliott V, Fernandez M, O’Neal L, et al. The REDCap Consortium: Building an International Community of Software Platform Partners. J Biomed Inform. 2019;95:103208.

29. Barton CJ, Rathleff MS. ‘Managing My Patellofemoral Pain’: the creation of an education leaflet for patients. Bmj Open Sport Exerc Medicine. 2016;2(1):e000086.

30. Burbach B, Barnason S, Thompson SA. Using “Think Aloud” to Capture Clinical Reasoning during Patient Simulation. Int J Nurs Educ Scholarsh. 2015;12(1):1–7.

31. Lundgrén-Laine H, Salanterä S. Think-Aloud Technique and Protocol Analysis in Clinical Decision-Making Research. Qual Health Res. 2010;20(4):565–75.

32. Swaine-Verdier A, Doward LC, Hagell P, Thorsen H, McKenna SP. Adapting Quality of Life Instruments. Value Health. 2004;7:S27–30.

33. Wolcott MD, Lobczowski NG. Using cognitive interviews and think-aloud protocols to understand thought processes. Curr Pharm Teach Learn. 2021;13(2):181–8.

34. Rathleff MS, Roos EM, Olesen JL, Rasmussen S. Exercise during school hours when added to patient education improves outcome for 2 years in adolescent patellofemoral pain: a cluster randomised trial. Brit J Sport Med. 2015;49(6):406.

35. Comins JD, Brodersen J, Siersma V, Jensen J, Hansen CF, Krogsgaard MR. How to develop a condition-specific PROM. Scand J Med Sci Spor. 2021;31(6):1216–24.

