## Supplemental material 1-8 for "Item generation for a new patient-reported outcome measure: The non-traumatic anterior knee pain (AKP)-YOUTH scale"

### APPENDIX 1. INTERVIEW GUIDE FOR IDENTIFICATIONS OF DOMAINS

| INTRODUCTION TO INTERVIEW | |
|---|---|
| **Information** | **Questions** |
| Thank you so much for wanting to be in an interview. | |
| Before we get started, I just want to tell you how it's going to happen. | |
| | • Along the way, I will ask you some questions. Remember that there is no answer that is wrong. I'm only interested in **your** experiences with knee pain, so everything you have to say is right. |
| Are you ready for us to get started on the first part? | |

| INTERVIEW BOX 1: ACTIVITIES OF DAILY LIVING AND KNEE PAIN | |
|---|---|
| **Information** | **Question** |
| Now I would like you to try to describe a completely ordinary day in your life, from the time you get up until you go to bed. | |
| (Interviewer describes ordinary day) | • I'll stop you right here. How do you experience your knees in the things you just described to me?<br>• Can you try to tell me more about it? |
| | • Is there anything during a normal day in your life that we forgot to talk about? |
| (Interviewer summarizes and makes the informant confirm or disagree) | |

| INTERVIEW BOX 2: LEISURE TIME AND KNEE PAIN | |
|---|---|
| **Information** | **Question** |
| Now we've talked a little bit about what a normal day in your life looks like and when you experience knee pain. Now I want to talk a little more about what you do in your spare time. | |
| | • Can you try to tell me what you do when you have free time?<br><br>• How do you experience your knees in the things you do when you have free time? |
| (Interviewer summarizes and makes the informant confirm or disagree) | |

| INTERVIEW BOX 3: SPORT AND KNEE PAIN | |
|---|---|
| **Information** | **Question** |
| Now I would like to hear a little about what kind of sport you do. | |
| | • Do you play sports?<br><br>• What kind of sport do you do?<br><br>• How do you experience your knees when you play sports? |
| (Interviewer summarizes and makes the informant confirm or disagree) | |

| INTERVIEW BOX 4: SOCIAL LIFE AND KNEE PAIN | |
|---|---|
| **Information** | **Question** |
| Now I want to hear a little bit about what you do when you are with your friends. | |
| | • What do you do when you are with your friends?<br><br>• How do you experience your knees in the things you do when you are with your friends? |
| (Interviewer summarizes and makes the informant confirm or disagree) | |

| INTERVIEW BOX 5: MENTAL WELLBEING AND KNEE PAIN | |
|---|---|
| **Information** | **Question** |
| Now I would like to hear a little about what it is like to have knee pain. | |
| | • What is it really like to have knee pain as a young person?<br><br>• Can you try to tell me more about it?<br><br>• How do your knees affect you in feeling good? |
| (Interviewer summarizes and makes the informant confirm or disagree) | |

| END OF INTERVIEW | |
|---|---|
| **Information** | **Question** |
| Now we've been through all the questions I had for you. I think you've been good at answering them. | |
| | • Is there anything you think we need to talk about? |

### APPENDIX 2: THINK-ALOUD INTERVIEW GUIDE

| DOMAIN(s)* | QUESTION(s) | NOTE |
|---|---|---|
| **Symptoms**<br><br>**Physical Activity**<br><br>**Social Activities**<br><br>**Emotional Impact** | Questions were framed as<br>"how do you understand…" and<br>"do you have other suggestions for…"<br><br>Ex. How do you understand the concept 'a typical week'?<br>- *Do you have other suggestions for framing?*<br><br>Furthermore, the questions were framed as "How do you think these questions cover the symptoms you experience?" and "do you think XYZ are redundant or difficult to understand?"<br><br>Adolescents were also asked about the timing of the questions. | Covers:<br>• Clarity,<br>• Framing<br>• Timing<br>• Relevancy |
| **End** | - Do you think that this questionnaire covers your experience with knee pain sufficiently?<br><br>- Other comments? | |

*This section was updated after step 2 and 3, but the interview guide was designed prior to the think-aloud test.

### APPENDIX 3. CHANGES BASED ON STEERING GROUP

| Original domains | New domains |
|---|---|
| Symptoms (e.g. stabbing, clicking etc.)<br>• Alterations in pain | Symptoms/Symptoms of pain |
| Limitations in school<br>(PA or recess) | |
| Limitation of daily living<br>(e.g. commute, sitting) | Limitations physical activity |
| Limitations of social activities (e.g. being with friends | Limitations of social activities (e.g. being with friends |
| Limitations in sports participation (e.g. playing their typical sport) E.g. opt-outs from sports. | |
| Ability to cope with knee pain (e.g. suppression of pain)<br>• (Psychological/self-manage pain with activity) | |
| Worries about the future<br>(What the knee can tolerate and how it affects the future) | |
| Emotions<br>(E.g. fear, frustration, sadness, rumination) | Emotional and/or psychological impact of pain. |
| Loss of belief in pain will improve | |
| Alterations in mood<br>(e.g., fluctuations, decrease) | |
| Exclusion from social circles<br>(Involuntary, voluntary) | |
| Sleep disturbance | |

***Red highlights changes**

****Arrows show how topics were formulated into new ones**

### APPENDIX 4. PRELIMINARY ITEM FORMULATION

| Domain | Suggestion for items |
|---|---|
| **Symptoms** | Try and think back to a typical week when answering these questions.<br><br>1. How often do you have knee pain?<br>Never<br>Rarely<br>Sometimes<br>Often<br>Always<br><br>2. How severe is your knee pain?<br>Not at all<br>Mild<br>Moderate<br>Severe<br>Extremely severe<br><br>3. Do you have difficulties with sleep due to your knee pain?<br>Never<br>Rarely<br>Sometimes<br>Often<br>Always |
| **Limitations in physical activity** | The next questions are about how your knee pain relates to physical activities. Try and think back to a typical week when you answer these questions.<br><br>4. How difficult do you find it to be physically active (for example, moving so quickly you become tired or out of breath) due to your knee pain?<br>Not at all difficult<br>A little bit difficult<br>Somewhat difficult<br>Very difficult<br>Extremely difficult<br><br>5. To what extent does your knee pain make it difficult for you to participate in sports or other activities that other kids/adolescents your age can do.<br>Not at all difficult<br>A little bit difficult<br>Somewhat difficult<br>Very difficult<br>Extremely difficult |

| | |
|---|---|
| | 6. How much do you avoid physical activities (sport, exercise or playing so you become tired or out of breath) due to your knee pain?<br>Not at all<br>A little bit<br>Somewhat<br>To a great degree<br>I cannot do these activities<br><br>How difficult do you find the following activities due to knee pain?<br><br>7. How difficult do you find it to sit with your knee bent? (Example: sitting in a chair for a prolonged period due to your knee pain)<br>Not at all difficult<br>A little bit difficult<br>Somewhat difficult<br>Very difficult<br>Extremely difficult<br><br>8. How difficult do you find it to change from prolonged sitting to standing due to your knee pain? (Example: from school class to recess)<br>Not at all difficult<br>A little bit difficult<br>Somewhat difficult<br>Very difficult<br>Extremely difficult<br><br>9. How difficult do you find it to take the stairs due to your knee pain?<br>Not at all difficult<br>A little bit difficult<br>Somewhat difficult<br>Very difficult<br>Extremely difficult<br><br>10. How difficult do you find it to ride a bike due to your knee pain?<br>Not at all difficult<br>A little bit difficult<br>Somewhat difficult<br>Very difficult<br>Extremely difficult<br>Not relevant for me |

| | |
|---|---|
| | 11. How difficult do you find it to run due to your knee pain?<br>Not at all difficult<br>A little bit difficult<br>Somewhat difficult<br>Very difficult<br>Extremely difficult<br><br>12. How difficult do you find it to jump due to your knee pain?<br>Not at all difficult<br>A little bit difficult<br>Somewhat difficult<br>Very difficult<br>Extremely difficult<br><br>13. How difficult do you find it to kneel due to your knee pain?<br>Not at all difficult<br>A little bit difficult<br>Somewhat difficult<br>Very difficult<br>Extremely difficult |
| **Limitations of social activities (e.g., being with friends)** | The next questions are about your knee pain when you are together with family and friends. Try and think back to a typical week for you when you answer these questions.<br><br>14. How difficult do you find it to do things with friends due to knee pain?<br>Not at all difficult<br>A little bit difficult<br>Somewhat difficult<br>Very difficult<br>Extremely difficult<br><br>15. How often do you feel that your knee pain limits you from doing things with friends?<br>Never<br>Rarely<br>Sometimes<br>Often<br>Always<br><br>16. How difficult do you find it to do things with your family due to knee pain?<br>Not at all difficult<br>A little bit difficult<br>Somewhat difficult |

| | |
|---|---|
| | Very difficult<br>Extremely difficult<br><br>17. How often do you feel that your knee pain limits you from doing things with your family?<br>Never<br>Rarely<br>Sometimes<br>Often<br>Always |
| **Emotional impact of pain.** | The next questions are about how your knee pain influences your mood and emotions. Try and think back to a typical week for you when you answer these questions.<br><br>18. To what degree do you experience that your knee pain makes it difficult to enjoy/have fun with your friends.<br>Not at all difficult<br>A little bit difficult<br>Somewhat difficult<br>Very difficult<br>Extremely difficult<br><br>19. How often do you experience your knee pain makes it difficult for you to have fun with your friends?<br><br>Never<br>Rarely<br>Sometimes<br>Often<br>Always<br><br>20. Do you become frustrated/angry about your knee pain?<br>Not at all frustrated/angry<br>A little bit frustrated/angry<br>Somewhat frustrated/angry<br>Very frustrated/angry<br>Extremely frustrated/angry<br><br>21. How often are you frustrated/ angry about your knee pain?<br>Never<br>Rarely<br>Sometimes<br>Often<br>Always<br><br>22. How sad/upset are you about your knee pain?<br>Not at all sad/upset<br>A little bit sad/upset |

| | |
|---|---|
| | Somewhat sad/upset<br>Very sad/upset<br>Extremely sad/upset<br><br>23. How often are you sad/upset with your knee pain?<br>Never<br>Rarely<br>Sometimes<br>Often<br>Always |

**APPENDIX 5a: DOMAINS, PRELIMINARY ITEMS, FEEDBACK AND CHANGES**

| ITEM | FEEDBACK | ACTIONS TAKEN | RATIONALE |
|---|---|---|---|
| **SYMPTOMS** | | | |
| How often do you have knee pain?<br><br>Never<br>Rarely<br>Sometimes<br>Often<br>Always | | | |
| How intense is your knee pain?<br><br>Not at all<br>Mild<br>Moderate<br>Severe<br>Extremely severe | Remove not at all<br><br>'Intense' may be difficult for adolescents to understand<br>Consider 'how severe' | No action was taken.<br><br>Change to how bad/severe and assess in the pilot. | The questionnaire is intended to capture changes over time and be used in studies where it is expected they may not have knee pain at follow-up.<br><br>"Intense" will be evaluated in the pilot to determine the most appropriate wording. |
| **LIMITATIONS IN PHYSICAL ACTIVITY** | | | |
| How difficult do you find it to sit with your knee bent for a longer period due to your knee pain?<br><br>Not at all difficult<br>A little bit difficult<br>Somewhat difficult<br>Very difficult<br>Extremely difficult | Sitting is not the only problem, but standing after sitting<br><br>Define longer period<br><br>Patients will have trouble with 'knee bend' | Added an additional item about standing after sitting<br><br>Quantify the time period for sitting in class<br><br>Add examples of' sitting in a chair' | Add a new question on standing up after they had sat with bend knees for a prolonged time |
| How difficult do you find it to take the stairs due to your knee pain? ^<br><br>Not at all difficult<br>A little bit difficult | | | |

| | | | |
|---|---|---|---|
| Somewhat difficult<br>Very difficult<br>Extremely difficult | | | |
| How difficult do you find it to ride your bike due to your knee pain?<br><br>Not at all difficult<br>A little bit difficult<br>Somewhat difficult<br>Very difficult<br>Extremely difficult | Not everyone will have a bike | Changed phrase to ride 'a' bike instead of 'your' bike<br><br>Add the N/A option to this and other items that are not applicable to all | Consider adding N/A to reflect that not all kids own a bike and then consider how such an item can be scored. |
| How difficult do you find it to run due to your knee pain?<br><br>Not at all difficult<br>A little bit difficult<br>Somewhat difficult<br>Very difficult<br>Extremely difficult | | | |
| How difficult do you find it to jump due to your knee pain?<br><br>Not at all difficult<br>A little bit difficult<br>Somewhat difficult<br>Very difficult<br>Extremely difficult | | | |
| **SOCIAL ACTIVITIES** | | | |
| How difficult do you find it to participate in social activities with your friends due to knee pain? (by social activities we mean doing things and being with friends). | All the questions are very alike and should be combined | None | We will further explore in pilot interviews to explore if we need to delete some questions (ie frequency may be difficult to answer as it may depend on how often you |

| | | | |
|---|---|---|---|
| Not at all difficult<br>A little bit difficult<br>Somewhat difficult<br>Very difficult<br>Extremely difficult | | | participate in that activity. |
| How often do you feel that your knee pain limits you from participating in social activities with your friends?<br><br>Never<br>Rarely<br>Sometimes<br>Often<br>Always | All the questions are very alike and should be combined | None | |
| How difficult do you find it to participate in social activities with your family due to knee pain?<br><br>Not at all difficult<br>A little bit difficult<br>Somewhat difficult<br>Very difficult<br>Extremely difficult | All the questions are very alike and should be combined | None | |
| How often do you feel that your knee pain limits you from participating in social activities with your family?<br><br>Never<br>Rarely<br>Sometimes<br>Often<br>Always | All the questions are very alike and should be combined | None | |
| How difficult do you find it to participate in social activities in school | q5 & q6:<br>What if patients are not studying but working? | Change to studying/working and a N/A option. | To make this applicable for the majority we will change the phrasing. |

| | | | |
|---|---|---|---|
| due to your knee pain?<br><br>Not at all difficult<br>A little bit difficult<br>Somewhat difficult<br>Very difficult<br>Extremely difficult | Change to studying/working and a N/A option | | |
| How often do you feel that your knee pain limits you from participating in social activities at school?<br><br>Never<br>Rarely<br>Sometimes<br>Often<br>Always | All the questions are very alike and should be combined<br><br>Adolescents may not understand social activities | None<br><br><br>Rephrasing to 'being with' | Changed due to a risk of adolescents not understanding the words. |
| **EMOTIONAL IMPACT OF PAIN** | | | |
| How difficult do you find it to have fun together with your friends due to your knee pain?<br><br>Not at all difficult<br>A little bit difficult<br>Somewhat difficult<br>Very difficult<br>Extremely difficult | The similarity between items for frequency & extent, suggestion to merge. | None | |
| How often is it difficult for you to have fun together with your friends due to your knee pain?<br><br>Never<br>Rarely<br>Sometimes<br>Often<br>Always | Suggestion to remove frequency category and go with extent of difficulty as previously discussed in group.<br><br>No action taken now<br><br>Consider timing after the pilot test. | None | See further up, frequency may cause problems in answering. |

| | | | |
|---|---|---|---|
| How often are you frustrated/ angry about your knee pain?<br><br>Never<br>Rarely<br>Sometimes<br>Often<br>Always | | | |
| How sad/upset are you about your knee pain?<br><br>Not at all sad/upset<br>A little bit sad/upset<br>Somewhat sad/upset<br>Very sad/upset<br>Extremely sad/upset | About sadness, need an extra question for Q3<br><br>Ex: "How frustrated/angry are you about your knee pain". | Add the extent answer option. | In the pilot work we need to explore how many different emotions we need to capture (or can capture from these young individuals) |
| How often are you sad/upset with your knee pain?<br><br>Never<br>Rarely<br>Sometimes<br>Often<br>Always | | | |

### APPENDIX 5b: DOMAINS, PRELIMINARY ITEMS, FEEDBACK AND CHANGES

| SUGGESTIONS OF ITEMS FOR OTHER DOMAINS | | | |
|---|---|---|---|
| **Item** | | **Action** | **Rationale** |
| Physical activity and sport | | Ad item on general physical activity "exercise/active" | |
| Pain location | I think you should include the pain location. I believe that patients with particular pain in/beneath the knee cap are much harder to treat than patients with more widespread or soft tissue pain, which could be important in respect to choose the right treatment and prognosis | No action taken. | This is not relevant for a PROM. |
| Could questions related to fear of pain and/or damage be added? | | No action taken. | This is beyond the scope of the PROM and may reflect more a trait rather than a state. |
| Sleep | Disturbed by knee pain, whether difficulty getting to sleep or waking with pain | Add in one item. "Does your knee pain influence your sleep?"<br><br>"Do you have difficulties with sleep due to your knee pain?" | Evidence suggest that some adolescent sleep worse due to pain. This is the reason for adding this question. |

### APPENDIX 6: CHANGES FOLLOWING THINK-ALOUD EXERCISE

| Domain | Suggestion for items |
|---|---|
| **Symptoms** | Try and think back to a normal week when answering these questions.<br><br>1. How often do you have knee pain?<br>Never<br>Almost Never<br>Sometimes<br>Often<br>Almost Always<br><br>2. How severe is your knee pain?<br>Not at all<br>Mild<br>Between mild and severe<br>Severe<br>Very severe<br><br>3. Do you have difficulties with sleep due to your knee pain?<br>Never<br>Almost Never<br>Sometimes<br>Often<br>Almost Always |
| **Limitations in physical activity** | The next questions are about how your knee pain relates to physical activities (sport, exercise or playing). Try and think back to a normal week when you answer these questions.<br><br>4. How difficult do you find it to be physically active due to your knee pain?<br>Not at all difficult<br>A little bit difficult<br>Somewhat difficult<br>Very difficult<br>Extremely difficult<br><br>**HELP**: Think about when you move so quickly you become tired or out of breath.<br><br>5. To what extent does your knee pain make it difficult for you to participate in sports or other activities that other kids your age can do.<br>Not at all difficult<br>A little bit difficult<br>Somewhat difficult |

| | |
|---|---|
| | Very difficult<br>Extremely difficult<br><br>6. How much do you avoid physical activities due to your knee pain?<br>I cannot do these activities<br>Not at all<br>A little bit<br>Somewhat<br>To a great degree<br><br>**HELP:** Think about sport, exercise or playing where you become tired or out of breath.<br><br>**How difficult do you find the following activities due to knee pain?**<br><br>7. To sit with your knee bent for longer periods?<br>Not at all difficult<br>A little bit difficult<br>Somewhat difficult<br>Very difficult<br>Extremely difficult<br><br>**HELP**: Think about sitting in a chair for an hour<br><br>8. To change from prolonged sitting to standing?<br>Not at all difficult<br>A little bit difficult<br>Somewhat difficult<br>Very difficult<br>Extremely difficult<br><br>**HELP:** Think about going from school class to recess<br><br>9. To take the stairs?<br>Not at all difficult<br>A little bit difficult<br>Somewhat difficult<br>Very difficult<br>Extremely difficult<br><br>10. To ride a bike?<br>Not at all difficult<br>A little bit difficult<br>Somewhat difficult<br>Very difficult<br>Extremely difficult |

| | |
|---|---|
| | Not relevant for me<br><br>11. To run?<br>Not at all difficult<br>A little bit difficult<br>Somewhat difficult<br>Very difficult<br>Extremely difficult<br><br>12. To jump?<br>Not at all difficult<br>A little bit difficult<br>Somewhat difficult<br>Very difficult<br>Extremely difficult<br><br>13. To kneel due to your knee pain?<br>Not at all difficult<br>A little bit difficult<br>Somewhat difficult<br>Very difficult<br>Extremely difficult |
| **Limitations of social activities (e.g., being with friends)** | The next questions are about your knee pain when you are together with family and friends. Try and think back to a normal week for you when you answer these questions. Be aware that in this section we will ask you "*how difficult"* and then "*how often"* meaning how often you will find it *that difficult*.<br><br>14. How difficult do you find it to do things with friends due to your knee pain?<br>Not at all difficult<br>A little bit difficult<br>Somewhat difficult<br>Very difficult<br>Extremely difficult<br><br>15. How often do you feel that your knee pain limits you from doing things with friends?<br>Never<br>Almost Never<br>Sometimes<br>Often<br>Almost Always<br><br>16. How difficult do you find it to do things with your family due to knee pain?<br>Not at all difficult<br>A little bit difficult |

| | |
|---|---|
| | Somewhat difficult<br>Very difficult<br>Extremely difficult<br><br>17. How often do you feel that your knee pain limits you from doing things with your family?<br>Never<br>Almost Never<br>Sometimes<br>Often<br>Almost Always |
| **Emotional impact of pain.** | The next questions are about how your knee pain influences your mood and emotions. Try and think back to a normal week for you when you answer these questions. Be aware that in this section we will ask you "*how difficult"* and then "*how often"* meaning how often you will find it *that difficult*.<br><br>18. To what degree do you experience that your knee pain makes it difficult to have fun with your friends.<br>Not at all difficult<br>A little bit difficult<br>Somewhat difficult<br>Very difficult<br>Extremely difficult<br><br>19. How often do you experience your knee pain makes it difficult for you to have fun with your friends?<br>Never<br>Almost Never<br>Sometimes<br>Often<br>Almost Always<br><br>20. Do you become annoyed about your knee pain?<br>Not at all annoying<br>A little bit annoying<br>Somewhat annoying<br>Very annoying<br>Extremely annoying<br><br>21. How often are you annoyed about your knee pain?<br>Never<br>Almost Never<br>Sometimes<br>Often<br>Almost Always<br><br>22. How sad are you about your knee pain?<br>Not at all sad |

| | |
|---|---|
| | A little bit sad<br>Somewhat sad<br>Very sad<br>Extremely sad<br><br>23. How often are you sad with your knee pain?<br>Never<br>Almost Never<br>Sometimes<br>Often<br>Almost Always |

### APPENDIX 7: ITEMS AND THEIR ORIGINS

| Domain | Suggestion for items: | The item origins from: |
|---|---|---|
| **Symptoms** | Try and think back to a typical week when answering these questions.<br><br>1. How often do you have knee pain?<br>Never<br>Rarely<br>Sometimes<br>Often<br>Always<br><br>2. How severe is your knee pain?<br>Not at all<br>Mild<br>Moderate<br>Severe<br>Extremely severe<br><br>3. Do you have difficulties with sleep due to your knee pain?<br>Never<br>Rarely<br>Sometimes<br>Often<br>Always | Inspired by the KOOS-Child |
| **Limitations in physical activity** | The next questions are about how your knee pain relates to physical activities. Try and think back to a | Inspired by "The Kidscreen" and McGill Pain Questionnaire |

| | | |
|---|---|---|
| | typical week when you answer these questions.<br><br>4. How difficult do you find it to be physically active (for example moving so quickly you become tired or out of breath) due to your knee pain?<br>Not at all difficult<br>A little bit difficult<br>Somewhat difficult<br>Very difficult<br>Extremely difficult<br><br>5. To what extent does your knee pain make it difficult for you to participate in sports or other activities that other kids/adolescents your age can do.<br>Not at all difficult<br>A little bit difficult<br>Somewhat difficult<br>Very difficult<br>Extremely difficult<br><br>6. How much do you avoid physical activities (sport, exercise or playing so you become tired or out of breath) due to your knee pain?<br>Not at all<br>A little bit<br>Somewhat | VAS/NRS Scale |

| | |
|---|---|
| | To a great degree<br>I cannot do these activities<br><br>How difficult do you find the following activities due to knee pain?<br><br>7. How difficult do you find it to sit with your knee bent e.g. sitting in a chair for a prolonged period due to your knee pain?<br>Not at all difficult<br>A little bit difficult<br>Somewhat difficult<br>Very difficult<br>Extremely difficult<br><br>8. How difficult do you find it to change from prolonged sitting to standing due to your knee pain? (Example: from school class to recess)<br>Not at all difficult<br>A little bit difficult<br>Somewhat difficult<br>Very difficult<br>Extremely difficult<br><br>9. How difficult do you find it to take the stairs due to your knee pain?<br>Not at all difficult<br>A little bit difficult<br>Somewhat difficult |

| | |
|---|---|
| | Very difficult<br><br>Extremely difficult<br><br>10. How difficult do you find it to ride a bike due to your knee pain?<br>Not at all difficult<br><br>A little bit difficult<br><br>Somewhat difficult<br><br>Very difficult<br><br>Extremely difficult<br><br>Not relevant for me<br><br>11. How difficult do you find it to run due to your knee pain?<br>Not at all difficult<br><br>A little bit difficult<br><br>Somewhat difficult<br><br>Very difficult<br><br>Extremely difficult<br><br>12. How difficult do you find it to jump due to your knee pain?<br>Not at all difficult<br><br>A little bit difficult<br><br>Somewhat difficult<br><br>Very difficult<br><br>Extremely difficult<br><br>13. How difficult do you find it to kneel due to your knee pain?<br>Not at all difficult |

| | |
|---|---|
| | A little bit difficult<br>Somewhat difficult<br>Very difficult<br>Extremely difficult |
| **Limitations of social activities (e.g. being with friends)** | The next questions are about your knee pain when you are together with family and friends. Try and think back to a typical week for you when you answer these questions.<br><br>14. How difficult do you find it to do things with friends due to knee pain?<br>Not at all difficult<br>A little bit difficult<br>Somewhat difficult<br>Very difficult<br>Extremely difficult<br><br>15. How often do you feel that your knee pain limits you from doing things with friends?<br>Never<br>Rarely<br>Sometimes<br>Often<br>Always<br><br>16. How difficult do you find it to do things with your family due to knee pain?<br>Not at all difficult<br>A little bit difficult<br>Somewhat difficult<br>Very difficult |

| | | |
|---|---|---|
| | Extremely difficult<br><br>17. How often do you feel that your knee pain limits you from doing things with your family?<br>Never<br>Rarely<br>Sometimes<br>Often<br>Always | |
| **Emotional impact of pain.** | The next questions are about how your knee pain influences your mood and emotions. Try and think back to a typical week for you when you answer these questions.<br><br>18. To what degree do you experience that your knee pain makes it difficult to enjoy/have fun with your friends.<br>Not at all difficult<br>A little bit difficult<br>Somewhat difficult<br>Very difficult<br>Extremely difficult<br><br>19. How often do you experience your knee pain makes it difficult for you to have fun with your friends?<br>Never<br>Rarely | Inspired by EQ-5D. |

| | |
|---|---|
| | Sometimes<br>Often<br>Always<br><br>20. Do you become frustrated/angry about your knee pain?<br>Not at all frustrated/angry<br>A little bit frustrated/angry<br>Somewhat frustrated/angry<br>Very frustrated/angry<br>Extremely frustrated/angry<br><br>21. How often are you frustrated/ angry about your knee pain?<br>Never<br>Rarely<br>Sometimes<br>Often<br>Always<br><br>22. How sad/upset are you about your knee pain?<br>Not at all sad/upset<br>A little bit sad/upset<br>Somewhat sad/upset<br>Very sad/upset<br>Extremely sad/upset<br><br>23. How often are you sad/upset with your knee pain? |

| | |
|---|---|
| | Never<br>Rarely<br>Sometimes<br>Often<br>Always |

**Appendix 8: Dual panel**

Fokus skal være på at spørgsmålene skal kunne forstås af aldersgruppen 10-19-årige.

| **Dansk** | | **Engelsk** | | **Comments or revisions** |
|---|---|---|---|---|
| **Domæne** | **Items** | **Domæne** | **Suggestion items** | **Domæne/ Items** |
| **Symptomer** | Prøv at tænke tilbage til den sidste uge, når du skal besvare disse spørgsmål.<br>1. Hvor ofte har du ondt i knæet?<br>Aldrig<br>Sjældent<br>Nogle gange<br>Ofte<br>Altid<br><br>2. Hvor slemme er dine knæsmerter?<br>Slet ikke<br>Milde<br>Moderat<br>Slemt<br>Ekstremt slemt<br><br>3. Har du problemer med at sove på grund af dine knæsmerter?<br>Aldrig<br>Sjældent<br>Nogle gange<br>Ofte<br>Altid | **Symptoms** | Think back to the last week when answering these questions.<br><br>1. How often do you have knee pain?<br>Never<br>Rarely<br>Sometimes<br>Often<br>Always<br><br>2. How severe is your knee pain?<br>Not at all<br>Mild<br>Moderate<br>Severe<br>Very severe<br><br>3. Do you have difficulties with sleep due to your knee pain?<br>Never<br>Rarely<br>Sometimes<br>Often<br>Always | |
| **Begrænsninger ved fysisk aktivitet.** | De næste spørgsmål handler om hvordan dine knæsmerter påvirker din fysiske aktivitet. Prøv at tænke tilbage til en typisk uge, når du skal besvare disse spørgsmål.<br><br>4. Hvor svært er det for dig at være fysisk aktiv på grund af dine knæsmerter? (for eksempel hvis du bevæger dig så hurtigt at du mister pusten eller bliver træt) | **Limitations in physical** | The next questions are about how your knee pain affects your physical activities (sport, exercise or playing). Think back to a normal week when you answer these questions.<br><br>4. How difficult is it for you to be physically active due to your knee pain? | |

| | | | |
|---|---|---|---|
| | Slet ikke svært<br>En lille smule svært<br>I nogen grad svært<br>Meget svært<br>Ekstremt svært<br><br>5. I hvor høj grad oplever du at knæsmerterne gør det svært for dig at deltage i sport og aktiviteter, som andre børn/unge i din alder laver/gør.<br>Slet ikke.<br>En lille smule svært.<br>I nogen grad svært.<br>Meget svært<br>Altid.<br><br>6. I hvor høj grad undgår du fysisk aktivitet (sport, træning eller leg som kan få dig til at miste pusten) på grund af dine knæsmerter?<br><br>Slet ikke<br>En smule<br>I nogen grad<br>I stor grad<br>Jeg kan ikke udføre disse aktiviteter<br><br>Hvor svært synes du det er at lave følgende aktiviteter:<br>7. Hvor svært synes du, at det er at sidde med knæene bøjet, fx sidde på en stol i længere tid på grund af dine knæsmerter?<br>Slet ikke svært<br>En smule svært<br>I nogen grad<br>Meget svært<br>Ekstremt svært<br><br>8. Hvor svært synes du, at det er at skifte position fra siddende i længere tid til at stå op på grund af dine | | HELP: Think about when you move so fast you become tired or out of breath.<br><br>Not at all difficult<br>A little bit difficult<br>Somewhat difficult<br>Very difficult<br>Extremely difficult<br><br>5. To what extent does your knee pain make it difficult for you to participate in sports or other activities that other kids your age can do.<br>Not at all difficult<br>A little bit difficult<br>Somewhat difficult<br>Very difficult<br>Extremely difficult<br>Not relevant for me<br><br>6. To what extent do you avoid physical activities due to your knee pain?<br>HELP: Think about sport, exercise or playing where you become tired or out of breath.<br><br>Not at all<br>A little bit<br>Somewhat<br>To a great degree<br>I cannot do these activities<br>Not relevant for me<br><br>How difficult do you find the following activities due to your knee pain?<br>7. To sit with your knee bent for longer periods?<br>HELP: Think about sitting in a chair for an hour<br>Not at all difficult<br>A little bit difficult<br>Somewhat difficult<br>Very difficult |

| | | | |
|---|---|---|---|
| | knæsmerter? (Fx efter at have siddet ned i en time inden frikvarter)<br>Slet ikke svært<br>En smule svært<br>I nogen grad<br>Meget svært<br>Ekstremt svært<br><br>9. Hvor svært synes du, at det er at gå på trapper på grund af dine knæsmerter?<br>Slet ikke svært<br>En smule svært<br>I nogen grad<br>Meget svært<br>Ekstremt svært<br><br>10. Hvor svært synes du, at det er at cykle på grund af dine knæsmerter?<br>Slet ikke svært<br>En smule svært<br>I nogen grad<br>Meget svært<br>Ekstremt svært<br>Ikke relevant for mig<br><br>11. Hvor svært synes du, at det er at løbe på grund af dine knæsmerter?<br>Slet ikke svært<br>En smule svært<br>I nogen grad<br>Meget svært<br>Ekstremt svært<br><br>12. Hvor svært synes du, at det er at hoppe på grund af dine knæsmerter?<br>Slet ikke svært<br>En smule svært<br>I nogen grad | | Extremely difficult<br>Not relevant for me<br><br>8. To change from prolonged sitting to standing?<br>HELP: Think about going from school class to recess<br>Not at all difficult<br>A little bit difficult<br>Somewhat difficult<br>Very difficult<br>Extremely difficult<br>Not relevant for me<br><br>9. To take the stairs?<br>Not at all difficult<br>A little bit difficult<br>Somewhat difficult<br>Very difficult<br>Extremely difficult<br>Not relevant for me<br><br>10. To ride a bike?<br>Not at all difficult<br>A little bit difficult<br>Somewhat difficult<br>Very difficult<br>Extremely difficult<br>Not relevant for me<br><br>11. To run?<br>Not at all difficult<br>A little bit difficult<br>Somewhat difficult<br>Very difficult |

| | | | |
|---|---|---|---|
| | Meget svært<br>Ekstremt svært<br><br>13. Hvor svært synes du, at det er at sidde på knæ på grund af dine knæsmerter?<br>Slet ikke svært<br>En smule svært<br>I nogen grad<br>Meget svært<br>Ekstremt svært | | Extremely difficult<br><br>12. To jump?<br>Not at all difficult<br>A little bit difficult<br>Somewhat difficult<br>Very difficult<br>Extremely difficult<br><br>13. To kneel?<br>Not at all difficult<br>A little bit difficult<br>Somewhat difficult<br>Very difficult<br>Extremely difficult |
| **Begrænsninger ved sociale aktiviteter (fx hænge ud med venner).** | De næste spørgsmål handler om dine knæsmerter når du er sammen med familie og venner. Prøv at tænke tilbage på en typisk uge, når du skal besvare disse spørgsmål. Vær opmærksom på, at vi i dette afsnit vil spørge dig både omkring hvor svært du oplever at noget er og efterfølgende hvor ofte det så er svært.<br><br>"hvor s og derefter "hvor ofte", hvilket betyder, hvor ofte du vil finde det så svært.<br><br>14. Hvor svært synes du det er, at lave ting med dine venner på grund af dine knæsmerter?<br>Slet ikke svært<br>En smule svært<br>I nogen grad<br>Meget svært<br>Ekstremt svært<br><br>15. Hor ofte føler du at dine knæsmerter begrænser/forhindrer dig i at lave ting med dine venner?<br>Aldrig | **Limitations of social activities (e.g., being with friends)** | The next questions are about your knee pain when you are together with family and friends. Try and think back to a normal week for you when you answer these questions. Be aware that in this section we will ask you both how difficult you find something to be and then how often, meaning how often you find it that difficult.<br><br>14. How difficult do you find it to do things with friends due to your knee pain?<br>Not at all difficult<br>A little bit difficult<br>Somewhat difficult<br>Very difficult<br>Extremely difficult<br><br>15. How often do you feel that your knee pain limits/prevents you from doing things with friends?<br>Never<br>Rarely<br>Sometimes<br>Often<br>Always |

| | | | |
|---|---|---|---|
| | Sjældent<br>Nogle gange<br>Ofte<br>Altid<br><br>16. Hvor svært synes du det er at lave ting med din familie på grund af dine knæsmerter?<br>Slet ikke svært<br>En smule svært<br>I nogen grad<br>Meget svært<br>Ekstremt svært<br><br>17. Hvor ofte føler du at dine knæsmerter begrænser dig i at lave ting med dine familie?<br>Aldrig<br>Sjældent<br>Nogle gange<br>Ofte<br>Altid | | 16. How difficult do you find it to do things with your family due to your knee pain?<br>Not at all difficult<br>A little bit difficult<br>Somewhat difficult<br>Very difficult<br>Extremely difficult<br><br>17. How often do you feel that your knee pain limits you from doing things with your family?<br>Never<br>Rarely<br>Sometimes<br>Often<br>Always |
| **Smerte og humør** | De næste spørgsmål handler om hvordan dine knæsmerter påvirker dit humør. Prøv at tænke tilbage til en typisk uge, når du skal besvare disse spørgsmål.<br><br>18. I hvilken grad oplever du at dine knæsmerter påvirker din evne til at nyde at være sammen med venner?<br>Slet ikke<br>En smule<br>I nogen grad<br>I stor grad<br>Jeg kan ikke være sammen med mine venner<br><br>19. Hvor ofte oplever du, at dine knæsmerter gør det svært at have det sjovt med dine venner?<br>Aldrig | **Emotionel impact of pain** | The next questions are about how your knee pain influences your mood and emotions. Try and think back to a normal week for you when you answer these questions.<br><br>18. To what degree do you experience that your knee pain makes it difficult for you to have fun with your friends.<br>Not at all difficult<br>A little bit difficult<br>Somewhat difficult<br>Very difficult<br>Extremely difficult<br><br>19. How often do you experience your knee pain makes it difficult for you to have fun with your friends?<br>Never<br>Rarely |

| | | | |
|---|---|---|---|
| | Sjældent<br>Nogle gange<br>Ofte<br>Altid<br><br>20. Er du frustreret (irriteret/sur) over dine knæsmerter?<br>Slet ikke frustreret/sur<br>En smule frustreret/sur<br>I nogen grad frustreret/sur<br>Meget frustreret/sur<br>Ekstremt frustreret/sur<br><br>21. Hvor ofte bliver du frustreret (irriteret/sur) over dine knæsmerter?<br>Aldrig<br>Sjældent<br>Nogle gange<br>Ofte<br>Altid<br><br>22. I hvilken grad bliver du ked af det over dine knæsmerter?<br>Slet ikke oprevet/ked af det<br>En smule oprevet/ked af det<br>I nogen grad oprevet/ked af det<br>Meget oprevet/ked af det<br>Ekstremt oprevet/ked af det<br><br>23. Hvor ofte bliver du ked af det over dine knæsmerter?<br>Aldrig<br>Sjældent<br>Nogle gange<br>Ofte<br>Altid | | Sometimes<br>Often<br>Always<br><br>20. How frustrated (annoyed/angry) are you with your knee pain?<br>Not at all frustrated<br>A little bit frustrated<br>Somewhat frustrated<br>Very frustrated<br>Extremely frustrated<br><br>21. How often are you frustrated (annoyed/angry) with your knee pain?<br>Never<br>Rarely<br>Sometimes<br>Often<br>Always<br><br>22. How sad are you about your knee pain?<br>Not at all sad<br>A little bit sad<br>Somewhat sad<br>Very sad<br>Extremely sad<br><br>23. How often are you sad about your knee pain?<br>Never<br>Rarely<br>Sometimes<br>Often<br>Always |
